# Early Identification of Hospital Visit Risk in Heart Failure Using Wearable-Derived Data

**DOI:** 10.64898/2026.03.26.26349411

**Authors:** Vedrana Ivezić, Jack Dawson, Rose Doherty, Sukanya Mohapatra, Mirna Issa, Shawn Chen, Gregg C Fonarow, Michael K Ong, William Speier, Corey W. Arnold

## Abstract

**Objectives:** Heart failure is a leading cause of mortality, necessitating identification of patients at increased risk needing intervention. In this study, we investigated if Fitbit data can reveal physiological trends associated with hospital visit risk.

**Materials and methods:** Individuals with heart failure (n=249) were randomized into three arms for prospective 180-day monitoring. All arms received a Fitbit and wireless weight scale. Arm 1 received devices only; Arm 2 received a mobile app with surveys; Arm 3 received the app plus financial incentives.

**Results:** 51 participants had hospital visits during the study period. These individuals took fewer steps (p=.002) and reported increased symptom severity (p=.044). Resting heart rate increased three days prior to a visit (p=.022). Baseline steps revealed a higher visit probability for less active participants (p=.003).

**Discussion and conclusion:** Passive physiological monitoring can effectively identify individuals at risk of health exacerbation, demonstrating the potential of wearable devices for timely clinical intervention.

## INTRODUCTION

Heart failure (HF) is a prevalent health condition with high mortality, morbidity, and cost to the healthcare system [1]. In the United States (U.S.), 6.7 million adults have HF, with the prevalence expected to increase to 10.3 million by 2040 [1]. HF is a leading cause of hospital admission in adults over the age of 65 [2], with hospitalizations responsible for the majority of around $31 billion annually spent on HF-related costs [3, 4]. With approximately a third of the U.S. population at risk of developing HF [1], the already substantial economic burden from this chronic disease will continue to increase [4].

Wearable technology can enable real-time monitoring and identification of patients at higher risk of hospitalization [5]. Non-invasive wearable technologies, such as external heart monitors, have previously demonstrated strong performance in predicting hospitalizations in people with HF [6, 7]. Specifically, a study using data from the All of Us research program [8] found that machine learning models using derived values from Fitbit watches could classify whether a patient with cardiovascular disease (including atrial fibrillation, coronary artery disease, myocardial infarction, cardiomyopathy, and heart failure) would have a hospitalization in the near future [9]. While heart failure is predominantly diagnosed in patients over 65 [10], this study investigated cardiovascular diseases in a notably younger cohort, with participant age capped at 66 years. Alerts from wearable-derived signals can also enable prompt intervention to reduce the probability of heart failure related hospitalizations [11]. While previous studies have primarily focused on younger age groups or specific heart related wearables, this study integrates Fitbit-derived metrics and patient reported symptom severity to understand risk in a population with an age distribution aligning with typical incidence age.

In this study, we evaluated Fitbit data collected from 249 participants over 180 days. We hypothesized that patterns in daily Fitbit measurements and self-reported symptom severity can identify participants at increased risk of hospitalizations and emergency department (ED) visits (together referred to as hospital visits). Daily Fitbit values and survey responses were evaluated using survival curve analysis and statistical comparisons to identify measurements correlated with increased hospital visit risk.

## METHODS

### Study Information

English-speaking adults aged 50-80 with an HF diagnosis identified using our institution’s electronic health record (EHR) were recruited for enrollment in our study between July 2021 and September 2024. Patients who owned a smartphone and met the eligibility criteria established through phone conversations and an initial engagement survey were enrolled in our 180-day study. The study was approved by the UCLA Institutional Review Board (IRB). Additional details regarding the study population design can be found in [12] and demographic details can be found in Table I.

**TABLE I.** Study population: Demographics of the 249 study participants who completed the 180-day study, organized by intervention group and category.

| Category | Sub-category | Group D<br>(n=72) | Group D+A<br>(n=89) | Group D+A+F<br>(n=88) |
| --- | --- | --- | --- | --- |
| Gender | Male | 46 (63.89%) | 58 (65.17%) | 57 (64.77%) |
|  | Female | 26 (36.11%) | 30 (33.71%) | 30 (34.09%) |
|  | Non-binary/third gender | 0 | 1 (1.12%) | 1 (1.14%) |
| Race | White | 52 (72.22%) | 72 (80.90%) | 65 (73.86%) |
|  | Black/African American | 7 (9.72%) | 5 (5.62%) | 8 (9.09%) |
|  | Asian | 6 (8.33%) | 5 (5.62%) | 7 (7.95%) |
|  | American Indian/Alaska Native | 1 (1.39%) | 0 | 1 (1.14%) |
|  | Native Hawaiian/Pacific Islander | 1 (1.39%) | 1 (1.12%) | 0 |
|  | More than one race | 3 (4.17%) | 1 (1.12%) | 5 (5.68%) |
|  | Don't know/refuse | 2 (2.78%) | 5 (5.62%) | 2 (2.27%) |
| Age | 50-59 | 15 (20.83%) | 21 (23.6%) | 21 (23.86%) |
|  | 60-69 | 22 (30.56%) | 32 (35.96%) | 35 (39.77%) |
|  | 70-79 | 35 (48.61%) | 30 (33.71%) | 29 (32.95%) |
|  | 80-89 | 0 | 6 (6.74%) | 3 (3.41%) |
| Education | High school | 6 (8.33%) | 3 (3.37%) | 4 (4.55%) |
|  | Some college/associate degree/trade school | 18 (25.00%) | 20 (22.47%) | 27 (30.68) |
|  | Bachelor's degree | 16 (22.22%) | 26 (29.21%) | 23 (26.14%) |
|  | Master's degree or above | 32 (44.44%) | 40 (44.94%) | 34 (38.64%) |
| Income | 0–25,000 | 8 (11.11%) | 8 (8.99%) | 5 (5.68%) |
|  | 25,001–50,000 | 8 (11.11%) | 6 (6.74%) | 12 (13.64%) |
|  | 50,001–75,000 | 5 (6.94%) | 8 (8.99%) | 11 (12.50%) |
| | \$75,001 or more | 46 (63.89%) | 62 (69.66%) | 52 (59.09%) |
|  | Don't know/refuse | 5 (6.94%) | 5 (5.62%) | 8 (9.09%) |
| Hispanic/Latino | No | 67 (93.06%) | 82 (92.13%) | 77 (87.5%) |
|  | Yes | 5 (6.94%) | 7 (7.87%) | 11 (12.5%) |
| Etiology | Ischemic | 40 (55.56%) | 53 (59.55%) | 45 (51.14%) |
|  | Non-ischemic | 24 (33.33%) | 22 (24.72%) | 25 (28.41%) |
|  | Do not have | 8 (11.11%) | 14 (15.73%) | 18 (20.45%) |
| LVEF median | 25-34 | 7 (9.72%) | 9 (10.11%) | 13 (14.77%) |
|  | 35-44 | 13 (18.06) | 15 (16.85%) | 12 (13.64%) |
|  | 45-54 | 15 (20.83%) | 10 (11.24%) | 16 (18.18%) |
|  | 55-64 | 20 (27.78%) | 22 (24.72%) | 17 (23.61%) |
|  | 65-75 | 6 (8.33%) | 9 (10.11%) | 7 (7.95%) |
|  | <25 | 2 (2.78%) | 3 (3.37%) | 7 (7.95%) |
|  | Do not have | 9 (12.50%) | 21 (23.60%) | 16 (18.18%) |
| NYHA | 1 | 6 (8.33%) | 13 (14.61%) | 13 (14.77%) |
|  | 2 | 25 (34.72%) | 33 (37.08%) | 23 (26.14%) |
|  | 3 | 7 (9.72%) | 6 (6.74%) | 12 (13.64%) |
|  | 4 | 0 | 0 | 3 (3.41%) |
|  | 1-2 | 2 (2.78%) | 1 (1.12%) | 1 (1.14%) |
|  | 2-3 | 4 (5.56%) | 6 (6.74%) | 8 (9.09%) |
|  | 3-4 | 2 (2.78%) | 0 | 3 (3.41%) |
|  | Do not have | 26 (36.11%) | 30 (33.71%) | 25 (28.41%) |

**TABLE II.** Classification results: Logistic regression classifiers fit on various feature combinations from the two week baseline period to predict whether a participant had a visit during the study. Best performance is in bold and second best is underlined.

| Feature(s) | Balanced Accuracy | AUROC | AUPRC |
| --- | --- | --- | --- |
| Steps | <u>0.641</u> | **0.671** | 0.273 |
| Heart rate | 0.550 | 0.442 | 0.136 |
| Symptoms | 0.571 | 0.525 | 0.313 |
| Heart rate + symptoms | 0.542 | 0.568 | 0.291 |
| Steps + heart rate | **0.642** | 0.651 | 0.310 |
| Steps + symptoms | 0.593 | <u>0.661</u> | <u>0.321</u> |
| Steps + heart rate + symptoms | 0.587 | 0.642 | **0.322** |

Participants were independently randomized into three groups, each receiving an activity tracker and a scale. Group D only received devices, Group D+A received devices and a mobile app, and Group D+A+F received devices and a mobile app with financial incentives based on their adherence during the study. Participants with an app received daily surveys with three questions including the impact of heart failure symptom severity on their daily life, adherence to their diet, and usage of prescribed medicines. Each question had three possible response options: “not really” (0), “moderately” (1), and “extremely” (2). Hospital visits, including ED visits and hospital admissions during the study period were also collected. Dates of the hospital visits were self-reported by participants during the study exit questionnaire or found through EHR chart review for those lost to follow up.

### Data analysis

Fitbit data for enrolled participants was retrieved via the Fitabase API service. Continuous data (e.g., steps) was first smoothed using a 2-day lagging rolling mean and then z-score normalized for each patient individually. Z-scores were calculated using a rolling mean and standard deviation, initialized from the first three nonmissing values and updated continuously.

We compared Fitbit data and survey responses between participants with hospital visits (Visit group) and those without (No Visit group) to identify predictors of hospital visits. Hospital visits were considered if they occurred after the third day of data collection, and participants were censored after their first visit.

Fitbit and survey data from the three week window prior to a visit were compared to random windows from the No Visit group with the Mann-Whitney U test. Random windows were sampled up to three weeks before a visit for each participant in the Visit group to evaluate if visit-related trends persisted throughout the study. The random windows selected prior to three weeks before a visit and the window prior to a visit were compared with the Wilcoxon signed-rank test.

Features with significant differences between visit and non-visit windows were evaluated using the first two weeks of data as a baseline to stratify hospital visit risk. Time-to-event (hospital visit) analysis was performed using Kaplan-Meier survival analysis, and the survival functions were compared using the log-rank test. Logistic regression evaluated the predictive performance of various feature combinations. Performance was evaluated using leave-one-out cross-validation and compared against random performance using a permutation test (n=1000). DeLong’s test was used to compare performance across models.

## RESULTS

The study group counts are shown in Figure 2. In 249 participants, we observed 80 hospital visits across 51 unique participants.

### Steps

Visit group participants had lower daily step counts than those with no hospital visits (Figure 1a). There was a statistically significant difference between the steps taken three weeks prior to a visit and randomly sampled three-week windows from the No Visit cohort (*p* = .002). Randomly sampled three-week windows from the Visit cohort compared to the random sampling from the No Visit cohort were also significantly different (*p* = .029). While not statistically significant (*p >* .1), there was a noticeable decreasing trend in steps beginning around two weeks prior to an event when compared to the randomly sampled windows from the Visit cohort.

**FIG. 1.**
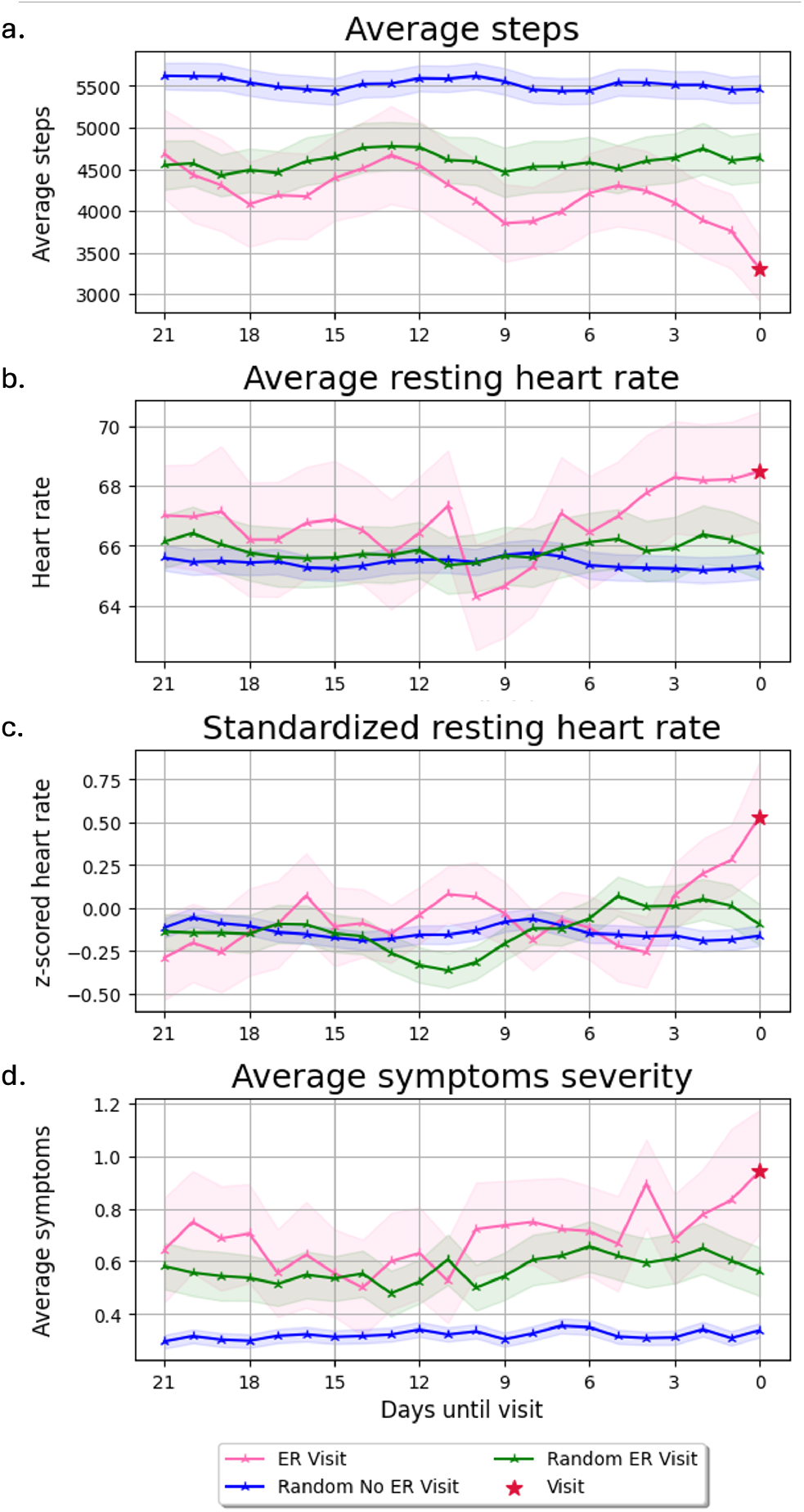
Differences between Visit and No Visit groups: Daily recorded values over three-week windows for steps, symptom severity, and resting heart rate. Three-week windows leading up to a visit (pink) and random sampled three-week windows prior to three weeks before a visit (green) were selected from the Visit cohort. Random three weeks were sampled from the entire study period from the No Visit cohort participants (blue). The hospital visit event is denoted by a red star.

**FIG. 2.**
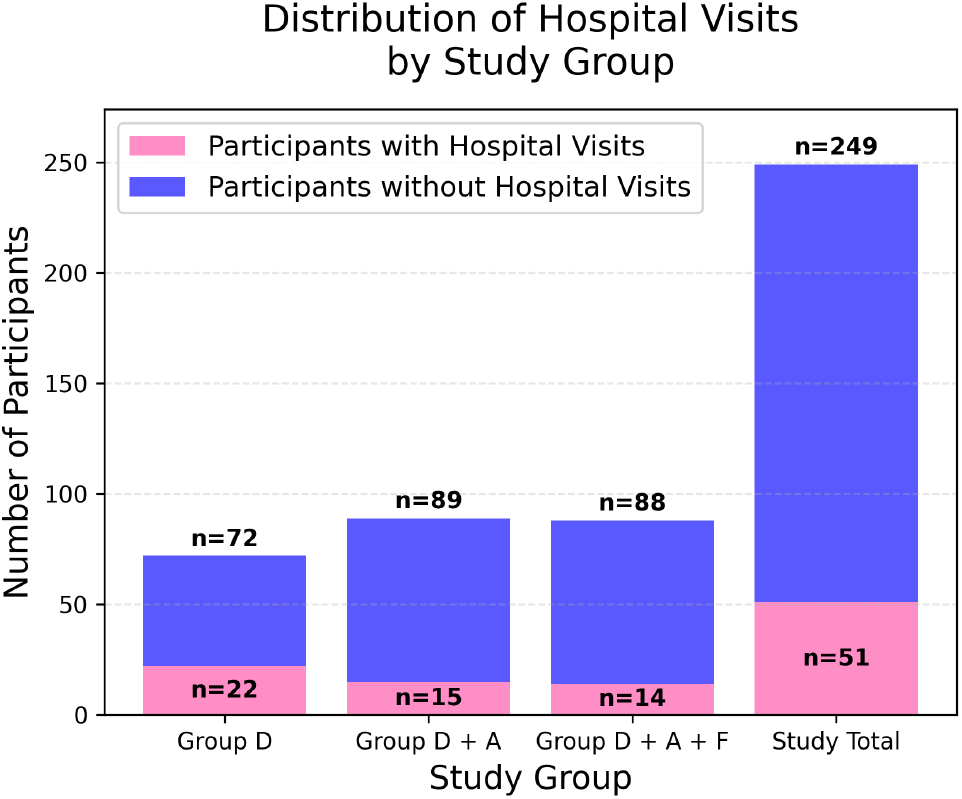
Distribution of hospital visits: Participants with at least one hospital visit during the 180-day study period are shown for the entire study population as well as stratified by study group.

Given the Visit cohort’s lower step counts, Ka-plan–Meier survival analysis was used to examine whether baseline activity levels were correlated with the time until a hospital visit (Figure 3a). The study cohort was split using the median average step count recorded during the first two weeks of data collection: participants with a lower average step count were high risk and those with higher step count were low risk. The high-risk group had a significantly higher probability of a hospital visit over time compared to the low-risk group (*p* = .003).

**FIG. 3.**
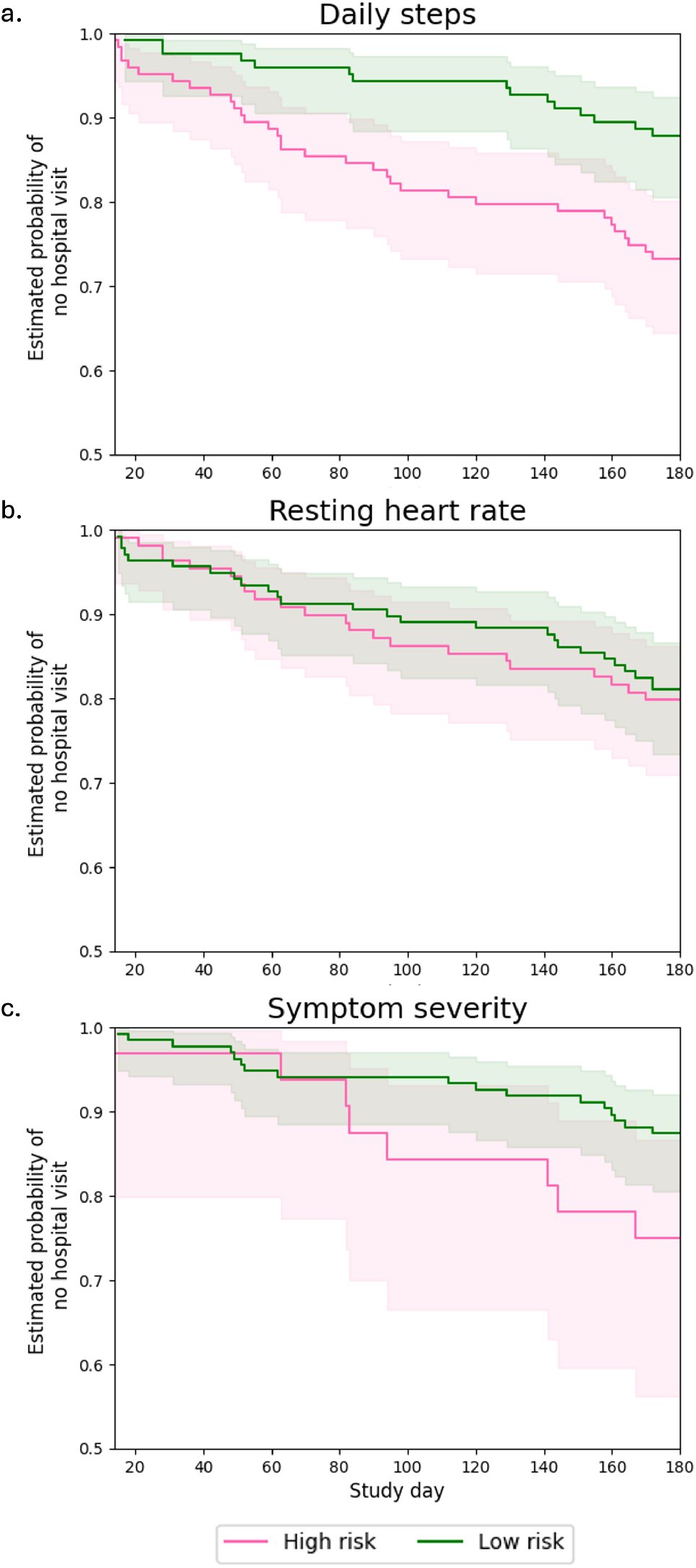
Survival curves: Participants were stratified into high (pink) and low (green) risk groups based on (a) median average steps, (b) average resting heart rate, and (c) average symptom severity of at least 1 during the first two weeks of data collection.

### Resting heart rate

Daily average resting heart rate increased during the three days preceding a visit compared to both prior days and the No Visit cohort (Figure 1b). Participants were stratified into high- and low-risk groups using the cohort’s mean two-week baseline resting heart rate as a threshold to evaluate whether resting heart rate is a predictor of exacerbation. Kaplan-Meier survival analysis found no significant difference (*p >* .78) between the high and low risk cohorts (Figure 3b).

To further evaluate these fluctuations over time while accounting for inter-subject variability, the average resting heart rate values were z-scored using each participant’s baseline values. There was no significant difference between the three weeks leading up to a visit compared to prior weeks for the Visit cohort. However, the normalized resting heart rate showed an increase three days prior to a visit (Figure 1c). Resting heart rate was significantly higher in the three days pre-visit compared to the preceding 18 days (*p* = .022) and the No Visit cohort (*p* = .022).

### Symptom severity

On average, participants in the Visit cohort reported higher symptom severity (Figure 1d) during the study than those in the No Visit cohort (*p* = .044). Symptom severity three weeks prior to a visit was significantly different from the No Visit cohort (*p* = 0.005) and randomly sampled three weeks from the Visit cohort (*p* = 0.011). An increase in symptoms three days prior to a visit was observed, but was not significantly different from randomly sampled three day windows (*p* = .0506) from the Visit group.

Kaplan-Meier analysis (Figure 3c) shows survival curves for high- and low-risk cohorts.

Participants were stratified as high-risk if average symptom severity was at least one during the two-week baseline; otherwise, they were considered low-risk. While there was an observed separation between cohorts, with the high-risk cohort having a higher probability of a hospital visit over time, the difference did not reach statistical significance (*p* = .073).

### Hospital visit prediction performance

Given that participants with hospital visits were less active and reported worse symptom severity throughout the study, we evaluated whether these features during only the two week baseline period could predict whether a participant would have a hospital visit during the remainder of the study. We fit logistic regression models with a balanced class weight using various combinations of daily steps, resting heart rate, and symptom severity from the first two weeks of the study to predict whether a participant would have a hospital visit during the remainder of the study. To allow for comparison with classification models using symptom severity, we dropped participants with no survey responses (Group D) from this part of the analysis.

Steps alone had the best performance of the univariate classifiers with an area under the receiver operating characteristic curve (AUROC) of 0.671. Classifiers excluding daily steps were not significantly different from random performance. Resting heart rate was not predictive alone but when used in combination with steps, the AUPRC improved modestly. When both symptoms and heart rate were combined with steps, the balanced accuracy and AUROC decreased compared to steps alone but the AUPRC increased.

## DISCUSSION

Daily Fitbit activity measurements and self-reported symptom severity were predictive of whether a participant would have a hospital visit in the near future. Participants with hospital visits generally took fewer daily steps, reported worse symptom severity, and had an increase in their resting heart rate three days before an event. When stratifying participants into low and high risk groups with data from a two week baseline period, average daily steps was the strongest risk separator. Daily steps were also the most important feature in classification models. Including heart rate and symptom information in the model with steps provided modest improvements, which suggests these variables may provide additional evidence for hospital visit prediction. Participants with visits which strongly correlated with higher symptom severity or an elevated resting heart rate were likely easier to identify when including these additional features. While AUPRC increased when adding symptoms and heart rate with the steps, the overall performance did not significantly improve. This finding suggests that passively collected activity data can characterize underlying health risk without the need for more invasive daily surveys.

Given that data from a two week baseline period could stratify hospital visit risk, future work should utilize these measurements as a baseline risk assessment to triage patients for increased monitoring and timely clinical intervention. Active surveillance following the baseline period should also explore monitoring fluctuations in resting heart rate which was found to increase shortly before a hospital visit. Early identification of at-risk patients could enable the targeted interventions necessary to decrease hospital visit rates and improve patient outcomes.

There are notable limitations of this work that should be addressed in subsequent studies. Measurements from wearable devices are inherently noisy, making low-intensity signals difficult to distinguish in limited sample sizes. There was also a significant difference between visit counts between groups D and D+A (p = .04) as well as between groups D and D+A+F (p=.03). The addition of an app, however, did not confound the trends identified in our results (see Supplementary Materials S2). As found in [12], the groups with an app were more adherent than group D which did not have an app, however, removing group D did not impact the findings of our work. In future studies, an app should be provided to all participants to increase engagement and reduce potential confounding variables.

## CONCLUSION

Wearable devices generate continuous signals reflecting an individual’s activity and physiological data. Our study provides additional evidence that these signals offer a new mode for monitoring a patient’s health at higher-frequency intervals relative to clinic visits. In the context of a chronic disease like HF, with high health and economic burden, wearables have the potential to bend the cost curve while improving health outcomes. Future studies should investigate pragmatic trials designed to realize these benefits.

## Data Availability

The data used in this study is available upon request.

## ACKNOWLEDGMENTS

This work is supported by a grant from the National Heart, Lung, and Blood Institute (R01HL141773).

## SUPPLEMENTARY MATERIALS

### Visit count differences between groups

A statistically significant difference was found in visit counts between the group with no app and those with an app (see Table S3). Figure S1 shows survival curves for each study group with group D having a sharper decline.

**TABLE S3.** Study Group Counts: Total number of participants and visits across study groups.

| Study Group | Participant Count | Participants with Visits | Total Visits |
| --- | --- | --- | --- |
| D | 72 | 22 (25%) | 34 |
| D+A | 89 | 15 (17%) | 18 |
| D+A+F | 88 | 14 (16%) | 28 |
| **Total** | **249** | **51 (20%)** | **80** |

### Addition of an app does not impact trends in steps

To ensure the use of an app was not a confounding variable in the step count analysis, we evaluated step survival curves stratified by group. Since both group D+A and D+A+F used apps we combined them in this analysis. Step count distribution during the baseline period of two weeks were not significantly different between groups (p=.67).

**FIG. S1.**
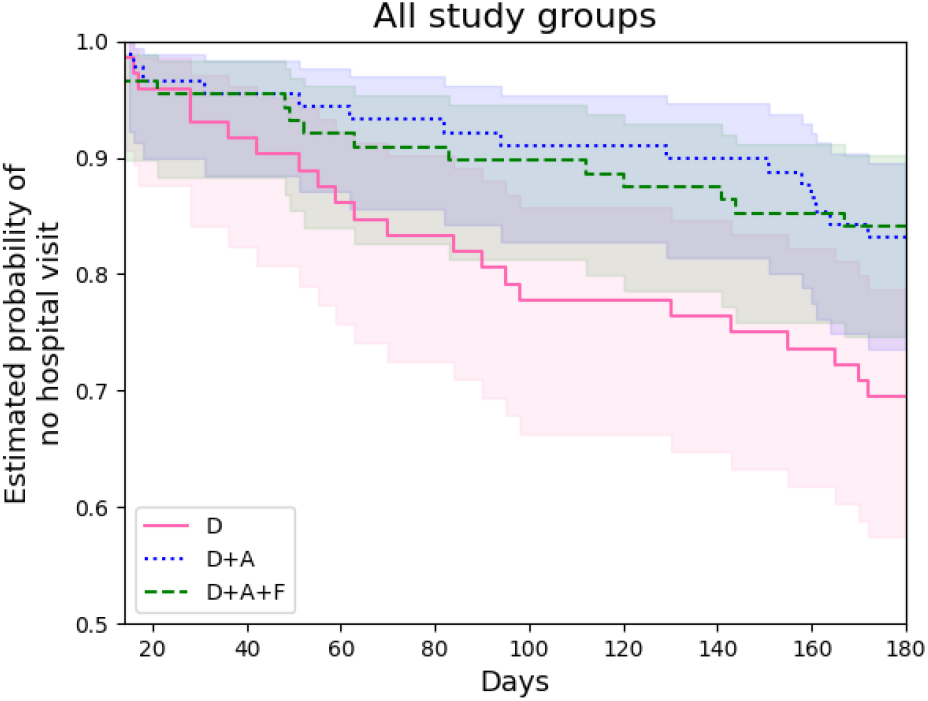
Kaplan-Meier curves by study group: Group D had significantly more visits than both Groups D+A and D+A+F.

**FIG. S2.**
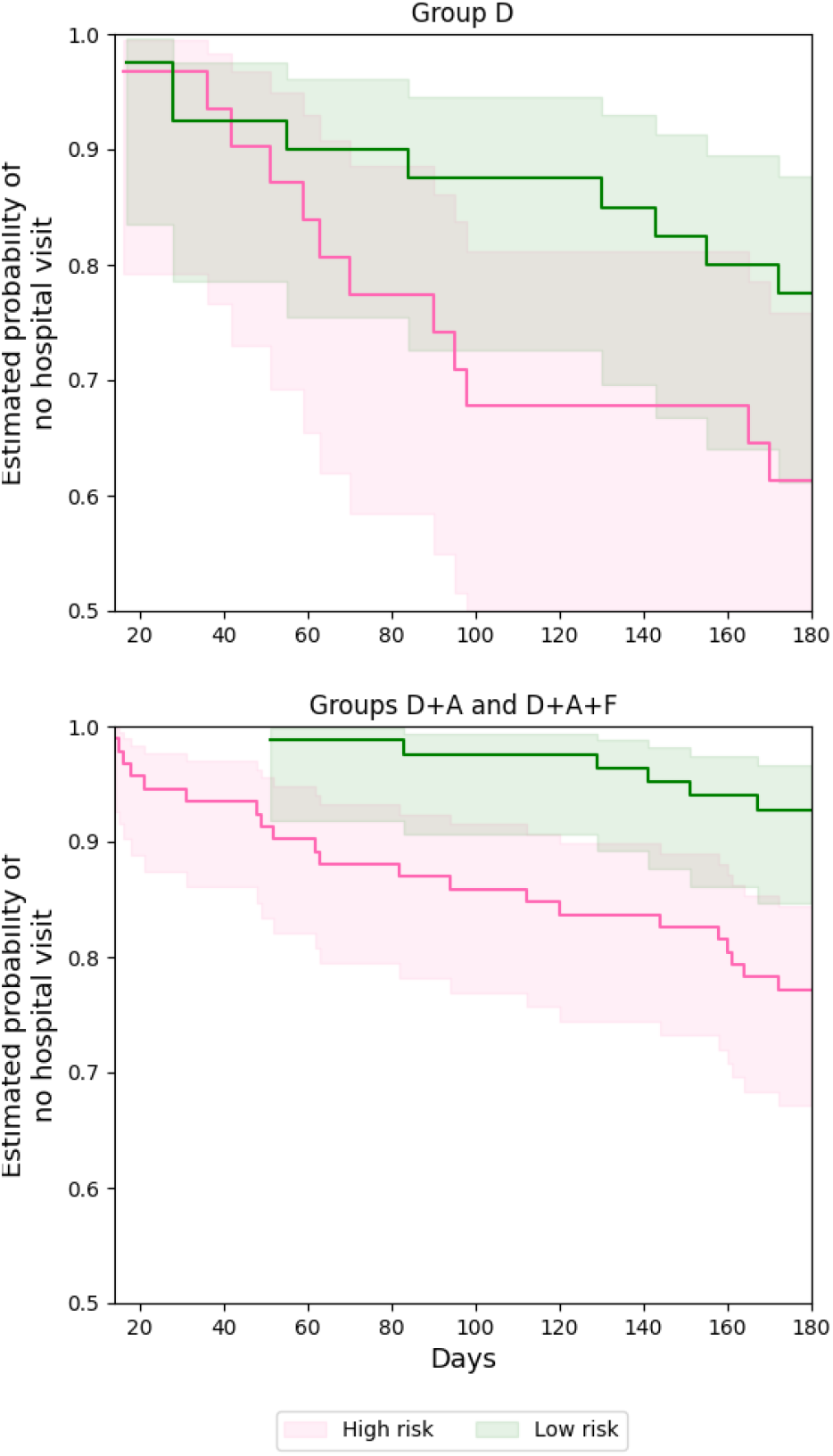
Group stratified Kaplan-Meier step curves: Steps were a predictor of hospital visits for all study group regardless of having an app.

